# Routine Xpert MTB/XDR for Tuberculosis Drug-Resistance Surveillance in Northern Morocco: A Four-Year Programmatic Analysis

**DOI:** 10.64898/2026.11.05.26364787

**Authors:** Ayoub Ez-Zari, Adil El Moussaoui, Abdennor Boulaich, Khalid Bouti, Zakaria Mennane, Noureddine Elmtili, Nadya Mezzoug

## Abstract

Drug-resistant tuberculosis surveillance requires diagnostic systems that can rapidly identify resistance beyond rifampicin and generate potentially actionable resistance information for routine public-health monitoring. We evaluated programmatic Xpert MTB/XDR use and characterized the molecular resistance profiles generated within a regional tuberculosis diagnostic network in northern Morocco. We retrospectively analyzed 258 Xpert MTB/XDR testing episodes performed from April 2022 to July 2026 and linked available Xpert MTB/RIF *Ultra* results with target-level molecular outputs. *Mycobacterium tuberculosis* complex was detected in 203 episodes (78.7%), of which 188 (92.6%) had an interpretable rifampicin result. Molecular rifampicin resistance was detected in 75/188 episodes (39.9%) and isoniazid resistance in 71/192 (37.0%); molecular multidrug-resistant tuberculosis was identified in 51/184 (27.7%) episodes with paired results. Among 186 episodes with interpretable rifampicin and additional resistance results, 75 (40.3%) had resistance to at least one additional drug class, including 21/112 (18.8%) rifampicin-susceptible and 54/74 (73.0%) rifampicin-resistant episodes. Among 92 resistant episodes with complete profiles, 66 (71.7%) showed resistance to two or more drug classes. *rpoB3-* and *rpoB4-*associated signals predominated among rifampicin-resistant episodes, *katG*-associated signals among isoniazid-resistant episodes, and *gyrA* targets among fluoroquinolone-resistant episodes. Mixed wild-type and mutation-associated signals occurred across *rpoB* and *gyrA*. These findings support the use of routine Xpert MTB/XDR testing as a component of regional drug-resistant tuberculosis surveillance, enabling systematic characterization of multiclass resistance and molecular resistance patterns within routine diagnostic networks. Such molecular surveillance should be integrated with phenotypic susceptibility testing and genomic sequencing to resolve discordant, atypical, or clinically consequential profiles and build a more comprehensive regional resistance-monitoring system.

## Introduction

Tuberculosis (TB), caused by members of the Mycobacterium tuberculosis complex (MTBC), remains a major global infectious disease threat. In 2024, an estimated 10.7 million people developed TB globally, while 8.3 million were newly diagnosed and notified (1). Despite progress in TB diagnosis and treatment, the global response remains off track to meet the End TB Strategy targets. Drug-resistant tuberculosis (DR-TB) remains a major challenge for effective treatment and disease control (1–3). Delayed detection of resistance can delay appropriate treatment, compromise patient management, and contribute to continued transmission. Rapid and comprehensive drug-susceptibility testing is therefore an important component of both clinical decision-making and DR-TB surveillance (4–6).

Conventional culture-based drug-susceptibility testing (pDST) remains important for comprehensive resistance assessment but is limited by prolonged turnaround times and substantial laboratory requirements. Rapid molecular diagnostics have consequently become central to the detection and surveillance of DR-TB, particularly in settings where timely resistance characterization is needed to guide treatment and strengthen programmatic monitoring (7).

Among the World Health Organization (WHO)-recommended molecular diagnostic tools, the GeneXpert platform is widely used for rapid detection of TB and drug resistance (8). Xpert MTB/RIF substantially reduced the time required to detect MTBC and rifampicin resistance, while Xpert MTB/XDR expanded molecular resistance testing to additional drug classes, including isoniazid, FQs, second-line injectable agents, and ethionamide. The assay interrogates resistance-associated targets including *katG, inhA, fabG1, gyrA, gyrB, rrs,* and *eis* sequences (8–10). This expanded molecular coverage can provide more comprehensive resistance information within routine diagnostic workflows and generate target-level data that may be useful for laboratory-based surveillance. However, Xpert MTB/XDR remains a targeted assay and does not capture all genetic mechanisms of resistance; consequently, it cannot replace comprehensive phenotypic DST or genomic characterization when broader resistance assessment is required (11, 12).

Beyond binary resistance classification, routine molecular diagnostics can generate target-associated profiles that describe the distribution and co-occurrence of resistance and provide additional information for molecular surveillance. Integrating rifampicin-associated *rpoB*profiles (Xpert MTB/RIF*Ultra*) testing with Xpert MTB/XDR data can therefore provide a multidimensional view of DR-TB within routine diagnostic networks. Such data may help characterize resistance patterns at the regional level and identify molecular signal variability that warrants further investigation, particularly in settings where systematic genomic surveillance remains limited (13, 14).

In Morocco, previous studies have identified recurrent resistance-associated variants in *rpoB, katG, inhA,* and *gyrA*, including established markers of rifampicin and isoniazid resistance (MDR-TB) (15–18). Studies from northern Morocco have also documented MDR-TB and recurrent *rpoB* profiles (19, 20). However, Moroccan evidence has largely been generated from selected clinical or reference-laboratory populations and has focused on line-probe assays, targeted sequencing, or other molecular approaches (21–27). Although Xpert MTB/XDR has been incorporated into Morocco’s 2024–2030 TB strategic plan for expanded resistance detection alongside phenotypic DST and sequencing (28), evidence from its routine programmatic use and the broader resistance information it generates remains limited.

We therefore conducted a retrospective study of routine Xpert MTB/XDR testing from its implementation in April 2022 through July 2026 within a regional TB diagnostic network in northern Morocco. We characterized programmatic testing outcomes and molecular resistance profiles, integrating rifampicin resistance and rpoB target data from linked Xpert MTB/RIFUltra results with Xpert MTB/XDR resistance-associated targets. We further assessed resistance combinations, target-level molecular patterns, and melting-temperature and melt-peak characteristics to evaluate the potential of routinely generated molecular data to strengthen regional DR-TB surveillance.

## Materials and Methods

### Study design, setting, and data sources

We conducted a retrospective, laboratory-based observational study of routine Xpert MTB/XDR testing at the Center of TB Diagnosis and Respiratory Diseases, Tétouan, Morocco. The laboratory served the Tangier-Tétouan-Al Hoceima (TTA) administrative region (northern Morocco), encompassing its eight prefectures and provinces **(Fig. 1)**. Xpert MTB/XDR was introduced in 2022 within the regional TB diagnostic network and was used for patients with rifampicin resistance or other predefined indicators of increased drug-resistance risk, including previous treatment or retreatment, known or suspected drug-resistant tuberculosis exposure, persistent acid-fast bacillus positivity after the fifth month of treatment, or the need for expanded resistance characterization to inform treatment planning. The retrospective dataset did not permit quantification of the indication-specific contribution of each referral criterion. All Xpert MTB/XDR assay records generated at the laboratory from April 2022 through July 2026 were eligible. No assay was excluded on the basis of final result. Target-level resistance analyses were restricted to MTB-detected episodes with interpretable results for the corresponding target/class.

**Figure 1.**
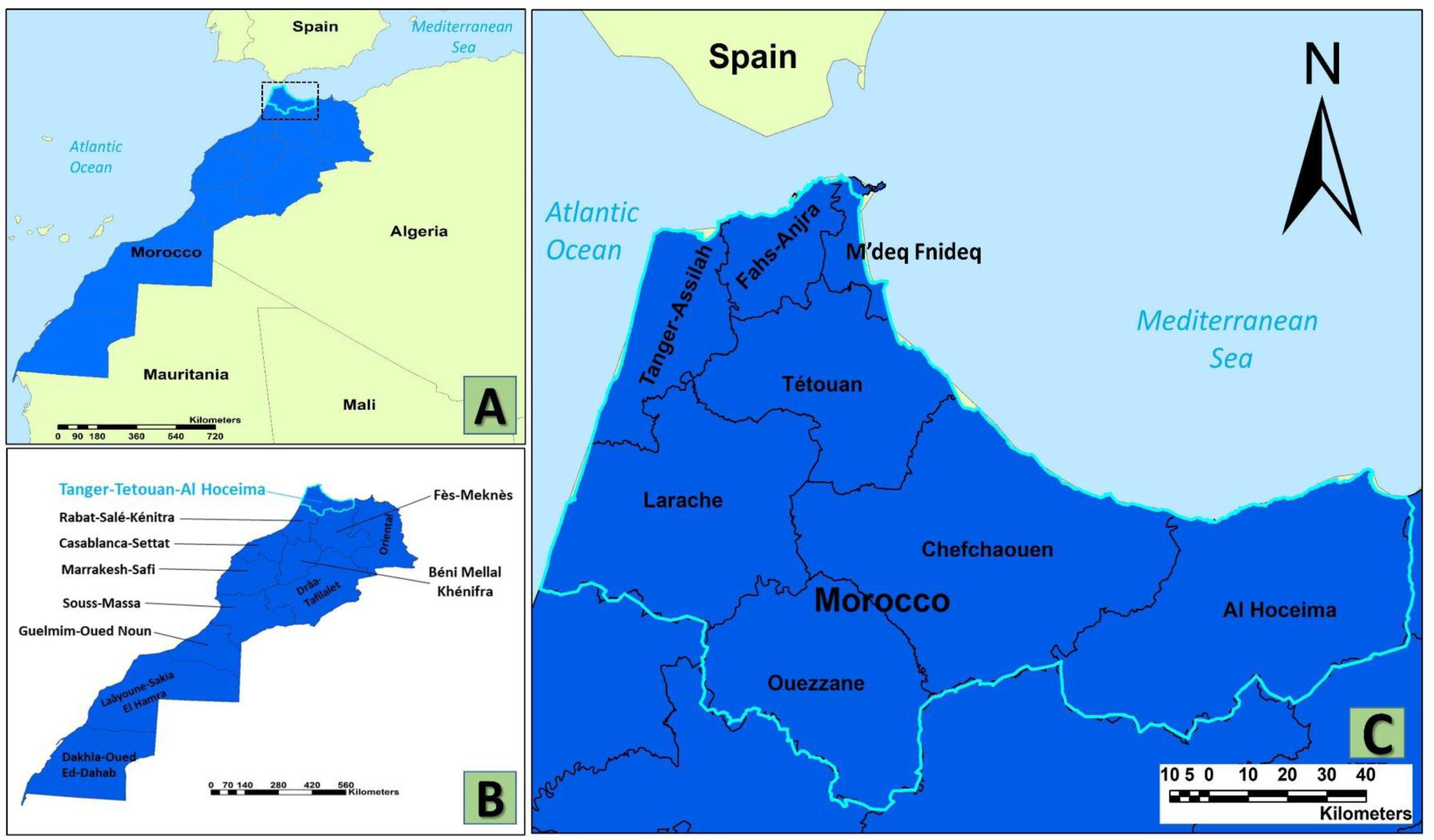
Geographic location of the study area: the Tangier-Tetouan-Al Hoceima region of northern Morocco. **(A)** Location of the Tangier-Tetouan-Al Hoceima (TTA) administrative region in Morocco, showing its proximity to Spain and the Mediterranean Sea. **(B)** Location of the TTA region within the national administrative division of Morocco. **(C)** Administrative structure of the TTA region, comprising the eight prefectures and provinces: Tanger-Assilah, Fahs-Anjra, M’diq-Fnideq, Tétouan, Larache, Chefchaouen, Ouezzane, and Al Hoceima. Maps were generated using ArcMap 10.8 from administrative boundary shapefiles obtained from SIG Maroc.

Because GeneXpert timestamps were inconsistently recorded, dates were used only for record linkage and identification of testing episodes, not temporal analyses. Xpert MTB/XDR records were linked, where available, to Xpert MTB/RIF-*Ultra* results using patient and sample identifiers. Age, sex, and referral address were obtained from routine laboratory registers. Each assay run was considered a testing episode; five repeat assays were performed after initially invalid results and yielded interpretable results, and these repeats were retained as separate episodes to characterize the programmatic testing pathway. For outcome-specific analyses, the subsequent interpretable result was used, whereas the preceding invalid assay was excluded from the corresponding denominator. Potential duplicates were assessed using patient, sample, and testing information.

### Xpert MTB/XDR and MTB/RIF testing

Xpert MTB/XDR and Xpert MTB/RIF-*Ultra* testing were performed according to the manufacturer’s instructions and routine procedures using the GeneXpert system (Cepheid, Sunnyvale, CA, USA) (28). Final interpretations and available target-level parameters, including cycle threshold (Ct), endpoint fluorescence (EndPt), amplification result, melting temperature (Tm), and melt-peak height, were retained. Targets included *inhA*, *katG*, *fabG1*, and *ahpC* for isoniazid resistance; *inhA* for ethionamide resistance; *gyrA1–3* and *gyrB2* for fluoroquinolones; and *rrs* and *eis* for second-line injectable resistance.

Linked Xpert MTB/RIF-*Ultra* records provided rifampicin results and available *rpoB1–4* data, including Ct, EndPt, amplification results, and wild-type- or mutation-associated Tm and melt-peak measurements. Melt-curve signals were treated as assay-level characteristics and were not interpreted as specific nucleotide substitutions without an established assay-specific interpretation.

### Resistance definitions and derived variables

Resistance classifications were based on final molecular assay interpretations. Molecular MDR-TB was defined as concurrent rifampicin and isoniazid resistance. Resistance variables were coded as detected, not detected, or indeterminate/missing. Resistance combinations were derived for rifampicin, isoniazid, fluoroquinolones, ethionamide, and second-line injectable agents (amikacin, kanamycin, and capreomycin).

Complete-profile analyses included episodes with interpretable results across all assessed resistance classes. The number of resistant drug classes and distinct resistance-associated molecular targets was calculated per episode; multiple signals within the same target were counted once.

### Molecular and programmatic analyses

All final Xpert MTB/XDR outcomes were retained, including MTB detected, MTB not detected, invalid, and no result. The primary denominator was all testing episodes; outcome-specific analyses used the relevant interpretable denominator, and non-interpretable results were not classified as susceptible.

Programmatic indicators included MTB detection, availability of linked rifampicin results, resistance by drug class, resistance combinations, and number of resistant classes. Molecular profiles integrated categorical resistance interpretations with target-level data, including resistance-associated targets and combined *rpoB*/Xpert MTB/XDR profiles.

For exploratory quantitative molecular analyses, all available numeric Tm and melt-peak measurements were retained and classified as WT or mutation-associated. Mixed WT/mutation signals were both retained. Thus, the unit of analysis was the assay-level molecular signal rather than the patient or testing episode. Tm and peak-height distributions were summarized as median (IQR) and compared between signal groups. Tm and melt-peak distributions were compared between wild-type and mutation-associated signal groups using non-parametric tests, as appropriate, without assigning specific nucleotide substitutions.

### Statistical analysis

Continuous variables were summarized as mean ± standard deviation or median and interquartile range (IQR), as appropriate, and categorical variables as frequencies and percentages. Fisher’s exact tests were used for sex comparisons, Mann–Whitney U tests for age and molecular signal distributions, and Monte Carlo tests based on Pearson χ² statistics for global province comparisons because of sparse cells. Odds ratios were calculated for male versus female comparisons where applicable.

Benjamini–Hochberg false-discovery-rate adjustment was applied across 18 prespecified demographic comparisons, and Holm adjustment across 10 Tm and 10 melt-peak comparisons. Two-sided *P* < 0.05 was considered statistically significant. Analyses were performed using IBM SPSS Statistics for Windows, version 26.0 (IBM Corp., Armonk, NY, USA).

### Missing data and quality control

Missing, invalid, no-result, and non-interpretable results were distinguished from negative results and were not imputed. When an invalid assay was repeated, the subsequent interpretable result was retained for the relevant analysis, while the initial assay remained classified as invalid. Analyses used the relevant interpretable denominator. Raw GeneXpert exports were reviewed for duplicates, formatting inconsistencies, missing values, and implausible measurements. Final interpretations were cross-checked against analyte-level results, and records with uncertain patient or specimen linkage were excluded from linkage-based analyses.

### Ethical considerations

The study used retrospectively collected routine laboratory and clinical data without additional intervention or specimen collection. Identifiable information was used only for internal linkage and removed from the analytical dataset. The protocol was approved by the Institutional Review Board of the Hospital-University Ethics Committee of Tangier (CEHUT; IRB No. AC112JV/2-025) and conducted in accordance with the Declaration of Helsinki. Informed consent was waived by the Institutional Review Board.

## Results

### 3.1 Study population and programmatic testing outcomes

A total of 258 Xpert MTB/XDR testing episodes were analyzed. Mycobacterium tuberculosis complex (MTBC) was detected in 203 (78.7%); 28 (10.9%) were MTBC-not-detected, 16 (6.2%) had no result, six (2.3%) were initially invalid, five of which were repeated and yielded interpretable results included among the 203 MTBC-detected episodes, and five (1.9%) lacked a final interpretation. Across all testing episodes, 201 (77.9%) involved men and 57 (22.1%) women; age was available for 253 episodes, with a median of 39 years (IQR, 30–49). Most specimens were sputum (94.2%), and referrals originated from seven prefecture/province categories, with Tétouan and Tangier accounting for 61.6% of all episodes. These distributions reflect the referral composition of the testing network rather than population-level estimates. A linked, interpretable rifampicin result was available for 188/203 MTB-detected episodes (92.6%), including 75 (39.9%) rifampicin-resistant episodes. The overall testing pathway is summarized in **Fig. 2**, and complete demographic, geographic, specimen, and testing-outcome characteristics are shown in **Table 1**.

**Figure 2.**
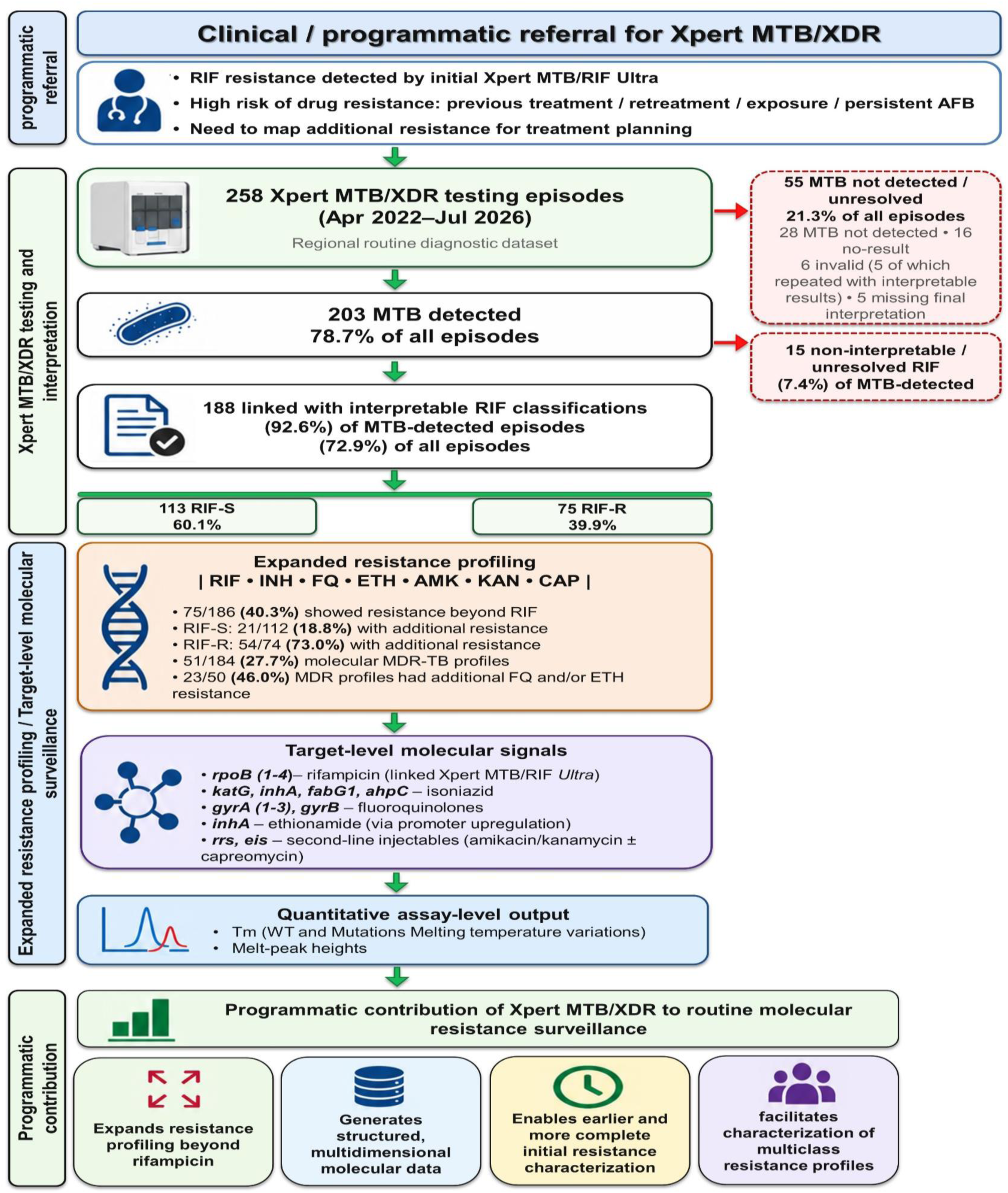
Programmatic pathway for Xpert MTB/XDR testing and molecular drug-resistance surveillance. Schematic representation of the programmatic pathway from clinical referral and Xpert MTB/XDR testing to expanded drug-resistance profiling and target-level molecular surveillance. The pathway includes the management of non-interpretable results and integration of assay-level molecular outputs, including melting temperature (Tm) and melt-peak signals, into routine resistance surveillance. Abbreviations: MTB, ***Mycobacterium tuberculosis***; RIF, rifampicin; INH, isoniazid; FQ, fluoroquinolones; ETH, ethionamide; MDR-TB, multidrug-resistant tuberculosis; Tm, melting temperature.

**Table 1.** Xpert MTB/XDR testing outcomes and characteristics of testing episodes (N=258) . The table summarizes the outcomes of all 258 Xpert MTB/XDR testing episodes and the demographic, geographic, and specimen characteristics of the tested population. Programmatic testing outcomes include MTB detection status and the availability of an interpretable linked RIF result. RIF susceptibility categories are reported among episodes with interpretable RIF results. Percentages are based on the relevant denominator indicated in the section heading.

| **Characteristic / programmatic indicator** | **n (%)** |
|---|---|
| **Programmatic testing outcomes (N=258)** |  |
| MTB detected | 203 (78.7) |
| MTB not detected | 28 (10.9) |
| No result | 16 (6.2) |
| Invalid | 6 (2.3) |
| Missing final interpretation | 5 (1.9) |
| **MTB detected with interpretable RIF result\*** | **188 (72.9)** |
| RIF susceptible | 113 (60.1) |
| RIF resistant | 75 (39.9) |
| **Characteristics of Xpert MTB/XDR testing episodes (N=258)** |  |
| **Sex** |  |
| Male | 201 (77.9) |
| Female | 57 (22.1) |
| **Age, years** |  |
| Available | 253 (98.1) |
| Mean ± SD | 41.4 ± 13.7 |
| Median (IQR) | 39 (30–49) |
| Range | 16–85 |
| <25 | 28 (10.9) |
| 25–34 | 52 (20.2) |
| 35–44 | 82 (31.8) |
| 45–54 | 54 (20.9) |
| 55–64 | 19 (7.4) |
| ≥65 | 18 (7.0) |
| **Prefecture/province of referral** |  |
| Tétouan | 94 (36.4) |
| Tangier | 65 (25.2) |
| Al Hoceima | 33 (12.8) |
| Fnideq | 25 (9.7) |
| Ouezzane | 21 (8.1) |
| Chefchaouen | 12 (4.7) |
| Larache | 8 (3.1) |
| **Specimen type** |  |
| Sputum | 243 (94.2) |
| Pus | 13 (5.0) |
| BAL | 1 (0.4) |
| Intestinal | 1 (0.4) |

\* RIF-susceptible and RIF-resistant episodes were counted among the 188 MTB-detected episodes with an interpretable linked RIF result. **Abbreviations:** BAL, bronchoalveolar lavage; IQR, interquartile range; MTB, *Mycobacterium tuberculosis* complex; RIF, rifampicin; SD, standard deviation.

### 3.2 Spectrum and architecture of drug resistance

Molecular resistance was detected most frequently for rifampicin (75/188, 39.9%) and isoniazid (71/192, 37.0%), followed by ethionamide (26/200, 13.0%) and fluoroquinolones (20/193, 10.4%). Amikacin, kanamycin, and capreomycin resistance were each detected in one episode; these calls occurred in the same episode and therefore represented a single episode with concurrent resistance to all three agents (**Table 2**).

**Table 2.**
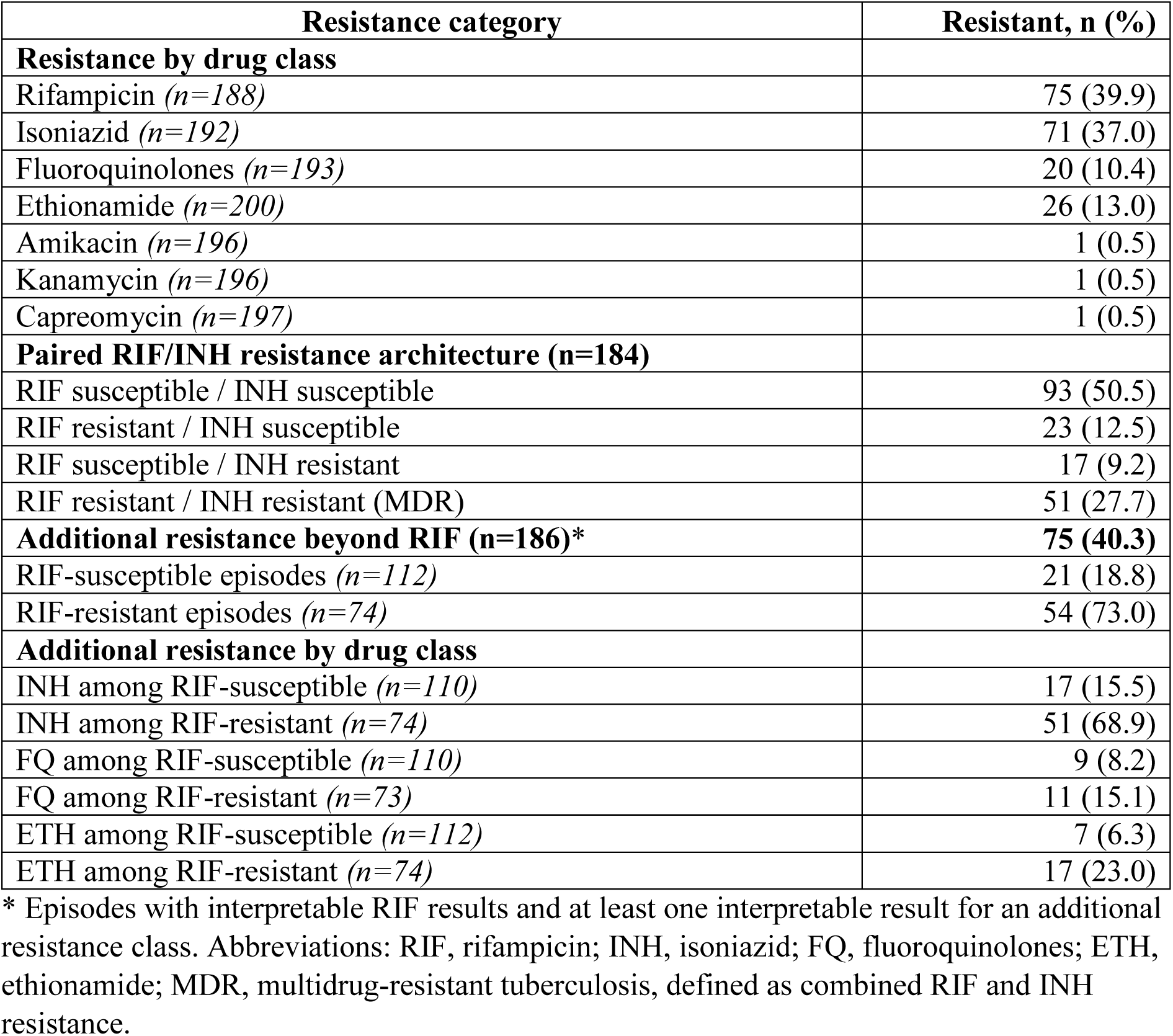
Distribution and architecture of drug resistance detected by Xpert MTB/XDR. ***n*** indicates the number of episodes with interpretable results for the corresponding resistance category. Resistance percentages were calculated using the indicated interpretable denominator. Additional resistance beyond RIF was defined as resistance to at least one additional assessed drug class among episodes with interpretable results. Non-interpretable, invalid, and missing results were excluded from the corresponding analyses.

Among 184 episodes with paired interpretable RIF and INH results, 51 (27.7%) showed concurrent resistance, corresponding to a molecular MDR-TB profile. Among 186 episodes with an interpretable rifampicin result and at least one additional resistance result, 75 (40.3%) had resistance to at least one additional drug class, including 18.8% of rifampicin-susceptible and 73.0% of rifampicin-resistant episodes.

Additional isoniazid resistance was strongly associated with rifampicin resistance (OR, 12.13; *P*<0.0001), as was ethionamide resistance (OR, 4.47; *P*=0.0014), whereas fluoroquinolone resistance was not significantly associated with rifampicin resistance (OR, 1.99; *P*=0.155) (**Table 3**).

**Table 3.**
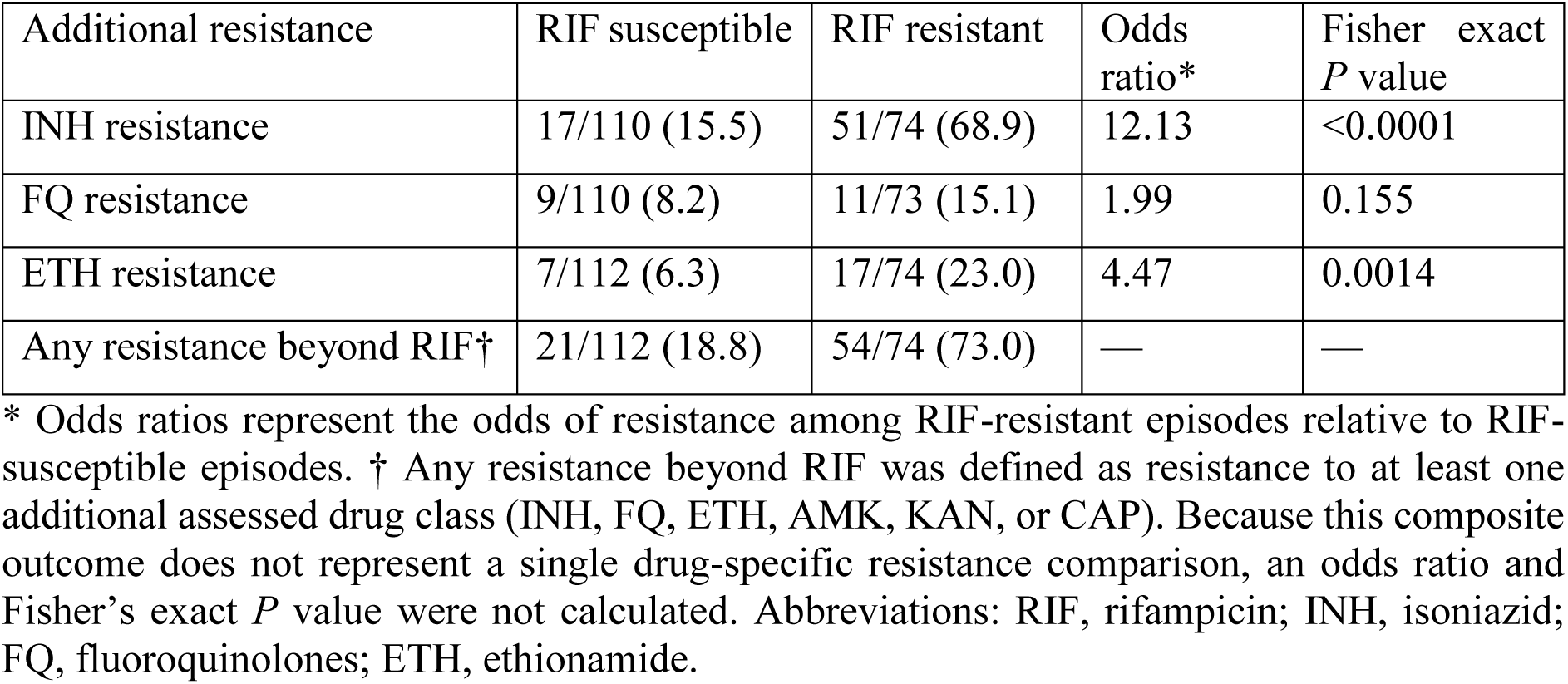
Additional molecular resistance according to rifampicin resistance status. Resistance frequencies are presented as *n/N (%)*, with denominators corresponding to episodes with interpretable results for the respective resistance category. Odds ratios compare the odds of additional resistance in RIF-resistant versus RIF-susceptible episodes. Fisher’s exact test was used to assess differences between RIF resistance groups.

### 3.3 Integrated resistance profiles

Among 180 episodes with complete interpretable profiles across all assessed drug classes (RIF, INH, FQ, ETH, AMK, KAN and CAP), 92 (51.1%) had resistance to at least one class and 66/92 (71.7%) had resistance to two or more classes. The most frequent patterns were RIF+INH (15.0%), RIF+INH+ETH (8.3%), and RIF+INH+FQ (3.9%). Among 50 molecular MDR-TB episodes with interpretable FQ and ETH results, 15 (30.0%) had MDR+ETH, seven (14.0%) MDR+FQ, and one (2.0%) MDR+FQ+ETH. FQ resistance was not significantly more frequent in MDR than non-MDR episodes (16.0% versus 9.0%; OR, 1.92; Fisher’s exact P=0.190), whereas ETH resistance occurred in 16/50 (32.0%) MDR episodes. Thus, the resistance burden was predominantly characterized by multiclass rather than single-class resistance (**Table 4**).

**Table 4.** Resistance-class profiles among episodes with complete interpretable results for all assessed drug classes (N=180). Resistance patterns represent the combination of drug classes with a resistant assay interpretation. “No detected resistance” indicates no resistance detected among the assessed drug classes. Resistance-class burden represents the total number of drug classes classified as resistant per episode; ≥2 indicates resistance to two or more drug classes. Percentages are calculated using the complete-profile denominator (N=180).

| **Resistance pattern** | **n** | **%** |
|---|---|---|
| Integrated resistance profiles |  |  |
| No detected resistance | 88 | 48.9 |
| RIF only | 19 | 10.6 |
| INH only | 4 | 2.2 |
| FQ only | 3 | 1.7 |
| RIF + FQ | 3 | 1.7 |
| INH + ETH | 7 | 3.9 |
| INH + FQ | 6 | 3.3 |
| RIF + INH (MDR only) | 27 | 15.0 |
| RIF + INH + ETH | 15 | 8.3 |
| RIF + INH + FQ | 7 | 3.9 |
| RIF + INH + FQ + ETH | 1 | 0.6 |
| Total | 180 | 100.0 |
| **Resistance-class burden** | **n** | **%** |
| 0 | 88 | 48.9 |
| 1 | 26 | 14.4 |
| 2 | 43 | 23.9 |
| 3 | 22 | 12.2 |
| 4 | 1 | 0.6 |
| ≥2 | 66 | 36.7 |

**RIF**, rifampicin; **INH**, isoniazid; **FQ**, fluoroquinolones; **ETH**, ethionamide; **MDR**, multidrug-resistant tuberculosis defined as combined RIF and INH resistance. Profiles represent mutually exclusive combinations of detected resistance among episodes with interpretable results for RIF, INH, FQ, ETH, AMK, KAN and CAP. The single episode with concurrent AMK, KAN and CAP resistance was not represented as an injectable-resistant combination because it did not have a complete interpretable profile across all assessed classes.

### 3.4 Demographic associations

Rifampicin resistance was associated with sex, occurring in 45.3% of men versus 20.0% of women (OR, 3.31; 95% CI, 1.43–7.66; *P*=0.0036; FDR-adjusted P=0.033), but not with age (*P*=0.211) or province (*P*=0.114). Isoniazid resistance showed no association with sex or age; an unadjusted provincial association (*P*=0.021) was not retained after false-discovery-rate adjustment (adjusted *P*=0.125). Ethionamide resistance varied significantly across provinces (*P*<0.001) and remained significant after adjustment (adjusted *P*=0.007), with the highest observed proportion in Chefchaouen (45.5%; 5/11). No association with sex or age was observed. FQ resistance, MDR profiles, and additional resistance beyond RIF showed no significant demographic associations (**Supplementary Table S4**).

Given the referral-based design and small provincial strata, these associations should be considered exploratory rather than population-level estimates.

### 3.5 Molecular resistance profiles

Target-level profiles differed by resistance class (**Table 5**). Among rifampicin-resistant episodes (n=75), *rpoB3* (46/75; 61.3%) and *rpoB4* (43/75; 57.3%) were the most frequently represented targets, and 45/75 (60.0%) involved at least two *rpoB* targets. Among isoniazid-resistant episodes (n=71), *katG* was the most frequent target (38/71; 53.5%), followed by *inhA* (26/71; 36.6%); single-target profiles predominated (64/71; 90.1%). *fabG1* was absent among the resistant episodes. All ethionamide-resistant episodes (n=26) had an *inhA*-associated signal, with 19 (73.1%) showing an *inhA*-only profile. Among fluoroquinolone-resistant episodes (n=20), *gyrA1* (18/20; 90.0%) and *gyrA3* (17/20; 85.0%) predominated, whereas no *gyrB2*-associated signal was retained. Seventeen episodes (85.0%) had signals involving at least two *gyrA* targets. The single episode with resistance to amikacin, kanamycin, and capreomycin had an *rrs*-associated signal without *eis*.

**Table 5.** Resistance-associated molecular profiles by drug class and target. Resistance-associated molecular target profiles detected across Xpert MTB/XDR and linked Xpert MTB/RIF Ultra results among episodes with resistance to each drug class.

| **Drug/class and Resistant episodes** | **Target/profile** | **n (%)** |
|---|---|---|
| **RIF(n=75)** | *rpoB1* | 15 (20.0) |
|  | *rpoB2* | 23 (30.7) |
|  | *rpoB3* | 46 (61.3) |
|  | *rpoB4* | 43 (57.3) |
|  | *≥2 rpoB targets* | 45 (60.0) |
|  | *rpoB3 + rpoB4* | 25 (33.3) |
| **INH (n=71)** | *katG* | 38 (53.5) |
|  | *inhA* | 26 (36.6) |
|  | *ahpC* | 14 (19.7) |
|  | *fabG1* | 0 |
|  | *katG only* | 31 (43.7) |
|  | *inhA only* | 19 (26.8) |
|  | *ahpC only* | 14 (19.7) |
|  | *inhA + katG* | 7 (9.9) |
|  | *≥2 INH-associated targets* | 7 (9.9) |
| **FQ (n=20)** | *gyrA1* | 18 (90.0) |
|  | *gyrA2* | 13 (65.0) |
|  | *gyrA3* | 17 (85.0) |
|  | *gyrB2* | 0 |
|  | *≥2 gyrA targets* | 17 (85.0) |
|  | *gyrA1 + gyrA2 + gyrA3* | 11 (55.0) |
|  | *gyrA1 + gyrA3* | 5 (25.0) |
| **ETH (n=26)** | *inhA-associated* | 26 (100) |
|  | *inhA only* | 19 (73.1) |
|  | *inhA + katG* | 7 (26.9) |
|  | *ahpC* | 0 |
|  | *fabG1* | 0 |
| **Injectables(n=1)** | *rrs* | 1 (100) |
|  | *eis* | 0 |

Values are n (%) among resistant episodes. Molecular targets are not mutually exclusive; percentages therefore may not sum to 100%. Target-associated signals represent assay-level results and should not be interpreted as definitive nucleotide-level mutation calls. RIF, rifampicin; INH, isoniazid; FQ, fluoroquinolone; ETH, ethionamide.

Mixed wild-type (WT) and mutation-associated signals were observed among rifampicin- and fluoroquinolone-associated targets. Among RIF-resistant episodes, 38 (50.7%) showed mixed signals in at least one *rpoB* target, involving *rpoB1* (9), *rpoB2* (15), *rpoB3* (15), and *rpoB4* (5); target-level counts were not mutually exclusive. Among FQ-resistant episodes, 3 (15.0%) showed mixed signals in *gyrA* targets, involving *gyrA1* (3), *gyrA2* (1), and *gyrA3* (2), with some episodes showing mixed signals at multiple *gyrA* targets. All episodes with mixed signals were classified as resistant according to the final assay interpretation. No mixed signals were observed for INH-, ETH-, or injectable-associated targets.

### 3.6 Quantitative molecular signatures

Mutation-associated signals showed distinct Tm distributions across the principal *inhA*, *katG*, *ahpC*, *rpoB*, and *gyrA* targets, with statistically significant differences across all analyzable comparisons (**Fig. 3; Supplementary Table S2**). Among INH-associated targets, median Tm values were lower for mutation-associated than WT signals for *inhA* (70.8 vs 76.2°C; Δ −5.4°C), *katG* (68.2 vs 73.7°C; Δ −5.5°C), and *ahpC* (66.0 vs 68.6°C; Δ −2.6°C; all Holm-adjusted *P*<0.001). Similar shifts were observed for *rpoB1–3* (Δ −3.8 to −2.7°C), whereas *rpoB4-A* showed an opposite shift (+6.5°C; *P*<0.001). Among FQ-associated targets, mutation-associated Tm values shifted downward for *gyrA1* (Δ −4.25°C; Holm-adjusted *P*<0.001) but upward for *gyrA2* (+5.4°C; *P*<0.001) and *gyrA3* (+5.2°C; *P*=0.004).

**Figure 3.**
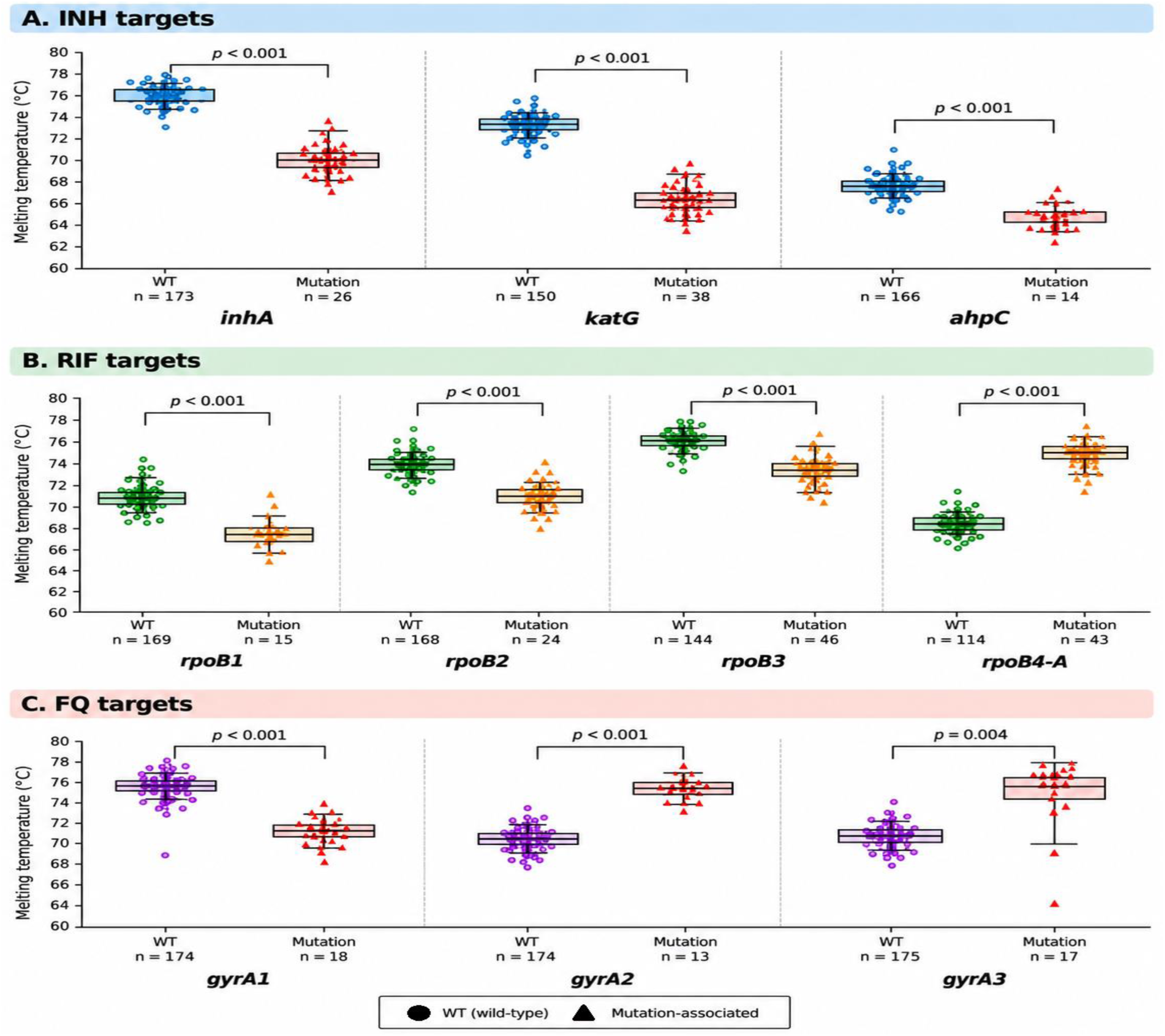
Melting-temperature distributions of wild-type and mutation-associated signals across selected resistance-associated targets. Box-and-scatter plots show melting temperature (Tm, °C) distributions for wild-type (WT) and mutation-associated signals across INH targets (A: *inhA, katG, ahpC*), RIF targets (B: *rpoB1, rpoB2, rpoB3, rpoB4-A*), and FQ targets (C: *gyrA1, gyrA2, gyrA3*). Each point represents an individual assay signal. Boxes indicate the median and interquartile range (IQR), with whiskers extending to 1.5 × IQR. The number of signals (*n*) is indicated below each group. Brackets indicate comparisons between WT and mutation-associated signals; P values are Holm-adjusted for multiple comparisons. The symbols in the legend denote WT (filled circles) and mutation-associated signals (filled triangles). RIF-associated rpoB1–4 signals were derived from linked Xpert MTB/RIF Ultra results.

Melt-peak heights also differed between WT and mutation-associated signals for several targets, but patterns were target-dependent and more variable. The largest increases were observed for *gyrA2* (median 64.2 versus 138.4) and *katG* (80.4 versus 121.0), whereas *rpoB3* showed substantially lower mutation-associated peak heights than wild-type signals (73.0 versus 37.5). *rpoB4-A* showed the opposite pattern, with higher mutation-associated peak heights (110.6 versus 54.0). These measurements were therefore treated as assay-level signal characteristics rather than specific biological effects or nucleotide substitutions (**Supplementary Table S3**).

## Discussion

The emergence of DR-TB has shifted the diagnostic challenge from MTBC detection to rapid resistance profiling for treatment and surveillance (29). However, molecular testing in many resource-limited settings remains focused on rifampicin, with broader profiling largely restricted to selected cohorts or research settings. In Morocco, resistance has similarly been characterized mainly through pDST, *rpoB* sequencing, and line-probe assays in selected populations (15, 16, 18–20, 27, 30, 31). In the present setting, Xpert MTB/XDR was introduced through a targeted workflow for patients with rifampicin resistance or other indicators of increased drug-resistance risk, including previous treatment or exposure and persistent acid-fast bacilli positivity after the fifth month of treatment.

To our knowledge, this is the first Moroccan study to characterize multidimensional resistance information from routine Xpert MTB/XDR.

Across 258 testing episodes over four years of implementation in northern Morocco, MTBC was detected in 203 (78.7%), while only a subset produced invalid, no-result, or non-interpretable outcomes. These findings highlight that the public-health value of molecular surveillance depends not only on resistance detection but also on specimen quality, result validity, repeat testing, and management of unresolved assays (32, 33). As a targeted assay within the diagnostic pathway, its effectiveness depends essentially on workflows that accommodate invalid, indeterminate, and repeat results (9, 34). Programmatic monitoring should therefore consider the complete diagnostic pathway rather than successful test outputs alone (35).

The expanded resistance panel identified additional resistance in 40.3% of episodes with interpretable rifampicin and additional resistance results, including 18.8% of rifampicin-susceptible and 73.0% of rifampicin-resistant episodes. This illustrates the value of broader molecular testing for identifying resistance that would not be captured by rifampicin-focused testing alone (7, 36, 37). Reflex Xpert MTB/XDR testing can provide a broader molecular resistance profile within a single workflow, complementing pDST and enabling more complete initial characterization.

The composition of additional resistance was drug-class specific. Isoniazid resistance was strongly associated with rifampicin resistance (OR, 12.13; p<0.0001), consistent with the molecular basis of MDR-TB (38). Ethionamide resistance was also enriched among rifampicin-resistant episodes (OR, 4.47; P=0.0014), whereas fluoroquinolone resistance did not differ significantly (OR, 1.99; P=0.155). Moroccan studies similarly report a substantial MDR burden among rifampicin-resistant isolates, but more heterogeneous second-line resistance across cohorts (19, 20, 30, 31, 39). These findings support rapid RIF–INH testing while highlighting the value of expanded testing for additional resistance that does not frequently follow the MDR backbone. Ethionamide results require caution because Xpert MTB/XDR does not capture the full genetic basis of phenotypic resistance (12).

Among episodes with complete resistance profiles, 71.7% of resistant episodes had resistance to two or more drug classes. Although these findings cannot be directly compared with studies based on selected MDR-TB cohorts or different testing methods (30, 31, 39), they indicate substantial multiclass resistance within the tested population. Under current WHO definitions, MDR/RR-TB with fluoroquinolone resistance is pre-XDR-TB, whereas XDR-TB additionally requires bedaquiline or linezolid resistance, neither assessed by Xpert MTB/XDR (36, 37). Thus, MDR+FQ profiles should not be classified as contemporary XDR-TB based on cartridge results alone (36).

The target-level findings further demonstrate how routine molecular data can add resolution to resistance surveillance. Rifampicin-resistant episodes frequently involved multiple rpoB targets, consistent with the concentration of resistance determinants within the rpoB rifampicin-resistance-determining region and patterns reported in Morocco (15, 16, 20, 40– 42). These profiles, however, represent assay-defined signatures rather than sequence-resolved genotypes. The absence of detectable targeted *rpoB* mutations in some phenotypically resistant isolates (16, 20) further highlights the potential contribution of off-target variants, disputed mutations and borderline resistance to molecular–phenotypic discordance (43–46). Isoniazid resistance was dominated by *katG-*and *inhA*-associated signals, with single-target profiles accounting for 90.1% of resistant episodes and no *fabG1* signal detected, consistent with the Moroccan reports of recurrent *katG315* and *inhA* promoter-associated variants (15, 18, 47–49). All ethionamide-resistant episodes carried an *inhA*-associated signal, although alternative mechanisms such as *ethA* and *ethR* are not captured by Xpert MTB/XDR (47, 50–53). The assay’s variable sensitivity for ethionamide resistance in multicenter evaluations further underscores this limitation (12). Fluoroquinolone resistance showed a predominantly *gyrA*-associated pattern, and no *gyrB* signal detected. This pattern is consistent with the recognized predominance of *gyrA* alterations and with previous Moroccan findings (30, 31, 39, 54–56). However, phenotypically resistant isolates without detectable *gyrA* or *gyrB* mutations have also been reported (30, 31, 39), indicating that targeted molecular testing may miss resistance arising through other mechanisms.

Mixed wild-type and mutation-associated signals occurred in *rpoB* and *gyrA*, while all corresponding episodes retained a resistant final interpretation. These patterns indicate molecular heterogeneity but cannot distinguish heteroresistance, mixed infection, or other within-sample processes by Xpert MTB/XDR alone (9, 58). Accordingly, raw assay outputs may provide information lost in binary resistance classification, but should not be interpreted as definitive evidence of heteroresistance; sequencing or other methods capable of resolving minority populations are required for further characterization (12, 59–61).

Raw melt-curve data revealed additional assay-level variation: Tm distributions differed clearly between wild-type and mutation-associated signals across the principal *rpoB*, *katG*, *inhA*, *ahpC*, and *gyrA* targets, consistent with the assay’s predefined Tm windows (8, 9). Peak heights were more target dependent and sensitive to bacillary load, specimen characteristics, and other technical factors (9, 62). Thus, Tm may support assay-level quality monitoring and interpretation of discordant or borderline results, whereas peak height is better regarded as a reaction-performance indicator than a specific resistance signature (7, 9, 41, 42).

Rifampicin resistance was more frequent among men, consistent with reports of male predominance in RR-TB in several settings (63). However, a global analysis of WHO surveillance data found no consistent association (64). Given the referral-based design and lack of adjustment for potential confounding, our finding should be considered exploratory rather than an independent association. Ethionamide resistance also varied across provinces, although sparse strata limit interpretation. These findings are therefore hypothesis-generating and warrant confirmation in population-based studies using adjusted analyses.

Together, these findings position Xpert MTB/XDR not only as a resistance diagnostic assay but as a potential component of regional DR-TB surveillance by generating resistance-class profiles, target-associated signatures, and quantitative molecular data within single routine workflows. International implementation studies similarly demonstrate its programmatic value while highlighting limitations related to specimen quality, heteroresistance, and incomplete target coverage (12, 32–34, 58, 65, 66). In northern Morocco, Broader deployment could provide more complete early resistance characterization, but cartridge costs, instrument capacity and health-system resources may constrain implementation (67). An interim approach would expand testing beyond rifampicin-resistant or otherwise high-risk patients to those with extensive radiographic disease, cavitation, persistent AFB positivity, severe or prolonged symptoms, or other broader indicators of high DR-TB risk (68).

A comprehensive regional surveillance framework should combine complementary laboratory approaches. Xpert MTB/RIF*Ultra* and Xpert MTB/XDR can provide rapid molecular resistance information, while phenotypic susceptibility testing and sequencing can resolve discordant, atypical, or clinically consequential profiles (7, 45, 69). Combining complementary molecular, phenotypic, and genomic approaches can broaden resistance detection beyond the targets covered by individual assays and help identify gaps in molecular target coverage, strengthening the evidence base for future diagnostic and surveillance strategies (10, 12, 14, 35, 45). Linking these diagnostic data with genotyping approaches such as MIRU-VNTR or whole-genome sequencing could further strengthen surveillance by helping distinguish transmitted from independently acquired drug resistance and identify potential resistant strain clusters (70–73). Such integration would move routine laboratory testing from isolated resistance detection toward a coordinated regional system for monitoring the distribution, diversity, and evolution of DR-TB.

This study has several limitations. Its retrospective, laboratory-based design reflects referral and testing selection rather than population-based sampling; therefore, estimates apply to the tested population rather than northern Morocco as a whole. Variable class-specific denominators also limited comparisons across resistance classes and inference on unique-patient burden. The absence of complete phenotypic and genomic reference data limited assessment of molecular–phenotypic discordance, off-target mechanisms, heteroresistance, and borderline resistance (45, 66, 74). The small number of injectable-resistant episodes and lack of treatment and clinical outcome data further limited interpretation. Thus, the study describes programmatic molecular resistance epidemiology rather than overall resistance prevalence, transmission dynamics, or treatment impact.

In conclusion, four years of routine Xpert MTB/XDR testing in northern Morocco demonstrate how expanded molecular diagnostics can transform routine laboratory testing into structured DR-TB surveillance data. By integrating rifampicin classification with additional resistance classes, multidrug profiles, target-associated signals, and quantitative assay features, routine diagnostic networks can generate broader information on the regional resistance landscape. The frequency of additional and multiclass resistance supports the value of broader resistance characterization, while the limitations of targeted assays highlight the need to integrate molecular, phenotypic, and genomic approaches. Such complementary strategies can strengthen resistance monitoring and provide a practical framework for developing more comprehensive DR-TB surveillance systems, particularly in settings where genomic surveillance remains limited.

## Acknowledgments

We acknowledge the Ministry of Health and Social Protection of Morocco for its institutional support and the staff of the Centre de Diagnostic de la Tuberculose et des Maladies Respiratoires (CDTMR), Tétouan, for their assistance with routine laboratory activities and data collection.

## Author contributions

A.E.Z.: Conceptualization, Methodology, Data curation, Formal analysis, Visualization, Writing – original draft, Writing – review & editing. A.E.M.: Investigation, Data curation, Validation, Writing – review & editing. A.B.: Investigation, Resources, Project administration, Writing – review & editing. K.B.: Conceptualization, Supervision, Methodology, Writing – review & editing. Z.M.: Investigation, Data curation, Validation, Writing – review & editing. N.E.: Investigation, Resources, Validation, Writing – review & editing. N.M.: Methodology, Supervision, Validation, Writing – review & editing. All authors approved the final version and agreed to be accountable for the work.

## Declaration of Generative AI Use

During the preparation of this manuscript, the authors used OpenAI ChatGPT (GPT-5.6 Luna) for language refinement, grammar and clarity checking, and identification of possible textual errors or inconsistencies. All suggestions were reviewed and verified by the authors, who retain full responsibility for the accuracy, originality, and integrity of the manuscript. ChatGPT was not used to generate research data, perform statistical analyses, or determine the study’s scientific findings or conclusions.

## Data availability

All data necessary to reproduce the analyses are provided in the manuscript and its supplemental material, including the complete assay-level dataset and codebook (Supplementary Data Set S1), melting-temperature analyses (Supplementary Table S2), melt-peak height analyses (Supplementary Table S3), and demographic associations (Supplementary Table S4).

## References

1. World Health Organization; 2025. Global tuberculosis report 2025. Geneva, Switzerland.

2. Zhang H, Guan W, Zhou J. 2025. Advances in the diagnosis of latent tuberculosis infection. Infect Drug Resist 18:483–493.

3. Pacchiarini N, Simkin F, Postans M, Ahern G, Song J, Brown C, Smith J, Williams C, Backx M, Thomas D. 2025. Genomic epidemiology of Mycobacterium tuberculosis in Wales. Sci Rep 15:31106.

4. World Health Organization. 2025. WHO consolidated guidelines on tuberculosis. Module 3: diagnosis. World Health Organization, Geneva, Switzerland.

5. World Health Organization. 2025. WHO consolidated operational handbook on tuberculosis. Module 4: treatment and care. World Health Organization, Geneva, Switzerland.

6. Farhat M, Cox H, Ghanem M, Denkinger CM, Rodrigues C, Abd El Aziz MS, Enkh-Amgalan H, Vambe D, Ugarte-Gil C, Furin J. 2024. Drug-resistant tuberculosis: a persistent global health concern. Nat Rev Microbiol 22:617–635.

7. Domínguez J, Boeree MJ, Cambau E, Chesov D, Conradie F, Cox V, Dheda K, Dudnyk A, Farhat MR, Gagneux S. 2023. Clinical implications of molecular drug resistance testing for Mycobacterium tuberculosis: a 2023 TBnet/RESIST-TB consensus statement. Lancet Infect Dis 23:e122–e137.

8. World Health Organization. 2021. Use of Xpert MTB/RIF and Xpert MTB/RIF Ultra on GeneXpert 10-colour instruments: WHO policy statement. World Health Organization, Geneva, Switzerland.

9. Cao Y, Parmar H, Gaur RL, Lieu D, Raghunath S, Via N, Battaglia S, Cirillo DM, Denkinger C, Georghiou S. 2021. Xpert MTB/XDR: a 10-color reflex assay suitable for point-of-care settings to detect isoniazid, fluoroquinolone, and second-line-injectable-drug resistance directly from Mycobacterium tuberculosis-positive sputum. J Clin Microbiol 59:e02314–20.

10. Penn-Nicholson A, Georghiou SB, Ciobanu N, Kazi M, Bhalla M, David A, Conradie F, Ruhwald M, Crudu V, Rodrigues C. 2022. Detection of isoniazid, fluoroquinolone, ethionamide, amikacin, kanamycin, and capreomycin resistance by the Xpert MTB/XDR assay: a cross-sectional multicentre diagnostic accuracy study. Lancet Infect Dis 22:242–249.

11. Sethi S, Sharma S, Aggarwal AN, Dhatwalia SK, Rana R, Yadav R. 2024. Xpert MTB/XDR assay: rapid TB drug resistance detection. Infection 52:2207–2212.

12. Massou F, Diarra B, Ba Diallo A, Bah KS, Vuchas C, Neh A, Sander M, Adebiyi EO, Aderemi BO, Agbla SC. 2026. Multicentre field evaluation of Xpert MTB/XDR in sub-Saharan Africa. ERJ Open Res 12:427–2025.

13. Reta MA, Maningi NE, Fourie PB. 2024. Patterns and profiles of drug resistance-conferring mutations in Mycobacterium tuberculosis genotypes isolated from tuberculosis-suspected attendees of spiritual holy water sites in Northwest Ethiopia. Front Public Health 12:1356826.

14. Georghiou SB, Tukvadze N, Rodrigues C, Omar SV, Cabibbe AM, Seifert M, Uplekar S, Ismail N, Ruhwald M, Suresh A. 2025. Targeted next-generation sequencing for drug-resistant tuberculosis diagnosis: implementation considerations for bacterial load, regimen selection and diagnostic algorithm placement. BMJ Glob Health 10:e019135.

15. Sabouni R, Kourout M, Chaoui I, Jordaan A, Akrim M, Victor TC, Maltouf Filali K, El Mzibri M, Lahlou O, El Aouad R. 2008. Molecular analysis of multidrug resistant Mycobacterium tuberculosis isolates from Morocco. Ann Microbiol 58:749–754.

16. Kourout M, Chaoui I, Sabouni R, Lahlou O, El Mzibri M, Jordaan A, Victor TC, Akrim M, El Aouad R. 2009. Molecular characterisation of rifampicin-resistant Mycobacterium tuberculosis strains from Morocco. Int J Tuberc Lung Dis 13:1440– 1442.

17. Chaoui I, Sabouni R, Kourout M, Jordaan AM, Lahlou O, Elouad R, Akrim M, Victor TC, Elmzibri M. 2009. Analysis of isoniazid, streptomycin and ethambutol resistance in Mycobacterium tuberculosis isolates from Morocco. J Infect Dev Ctries 3:278–284.

18. Ennassiri W, Jaouhari S, Sabouni R, Cherki W, Charof R, Filali-Maltouf A, Lahlou O. 2018. Analysis of isoniazid and rifampicin resistance in Mycobacterium tuberculosis isolates in Morocco using GenoType® MTBDRplus assay. J Glob Antimicrob Resist 12:197–201.

19. Karimi H, En-Nanai L, Oudghiri A, Chaoui I, Laglaoui A, Bourkadi JE, El Mzibri M, Abid M. 2018. Performance of GenoType® MTBDRplus assay in the diagnosis of drug-resistant tuberculosis in Tangier, Morocco. J Glob Antimicrob Resist 12:63–67.

20. Karimi H, Oudghiri A, En-Nanei L, Mzibri M El, Laglaoui A, Chaoui I, Abid M. 2020. Frequency of genomic mutations mediating resistance of Mycobacterium tuberculosis isolates to rifampicin in Northern Morocco. Rev Inst Med Trop Sao Paulo 62:e37.

21. Bennani K, Bukassa G, Khattabi A, Akrim M, Maaroufi A. 2018. Evaluation of drug-resistant tuberculosis surveillance system before and after implementing Genexpert in Morocco, 2007-2016. Iproceedings 4:e10535.

22. Hanane M, Hanane Z, Lamiae A, Youssef K, Mouhcine M, Said Z. 2020. GeneXpert’s performance in the diagnosis of pulmonary and extrapulmonary tuberculosis. Am J Lab Med 5:14–17.

23. Benaissa E, Benlahlou Y, Bssaibis F, Maleb A, Elouennass M. 2022. Evaluation of a Molecular Test for Detection of Mycobacterium tuberculosis Isolates Resistant to Rifampicin and Isoniazid. Clin Lab 68:532.

24. Mechal Y, Benaissa E, El Mrimar N, Benlahlou Y, Bssaibis F, Zegmout A, Chadli M, Malik YS, Touil N, Abid A. 2019. Evaluation of GeneXpert MTB/RIF system performances in the diagnosis of extrapulmonary tuberculosis. BMC Infect Dis 19:1069.

25. Aainouss A, Gh M, Belghiti A, Bennani K, Lamaammal A, Chetioui F, Messaoudi M, Blaghen M, Mouslim J, Khyatti M. 2022. Performance of GeneXpert MTB/RIF in the diagnosis of extrapulmonary tuberculosis in Morocco. Инфекция и иммунитет 12:78– 84.

26. Maafi K, Nafi I, Abidar H, Laariche F, Nabil M, Souly K, Aboudkhil S. 2025. Diagnostic Performance of GeneXpert MTB/RIF for Pulmonary and Extra-Pulmonary Tuberculosis in Morocco. Natural Built Social Environment Health 1:6–42.

27. Laatri S, El Kassimi S, Bentaleb EM, El Messaoudi MD, Irobi J, Belkadi B, Filali-Maltouf A, Ait Benhassou H. 2025. Characterization of drug resistance mutations in Mycobacterium tuberculosis isolates from Moroccan patients using deeplex targeted next-generation sequencing. Microorganisms 13:2163.

28. Ministry of health and social protection Morocco. 2024. National strategic plan for tuberculosis prevention and control, Morocco 2024-2030. Rabat.

29. Cepheid package insert 2021. 2021. Cepheid. Xpert MTB/XDR package insert. Document No. 302-3514. https://www.cepheid.com/content/dam/www-cepheid-com/documents/package-insert-files/Xpert%20MTB-XDR%20ENGLISH%20Package%20Insert%20302-3514%20Rev%20C.pdf. Retrieved 15 September 2026.

30. Tiberi S, Utjesanovic N, Galvin J, Centis R, D’Ambrosio L, van den Boom M, Zumla A, Migliori GB. 2022. Drug resistant TB–latest developments in epidemiology, diagnostics and management. International Journal of Infectious Diseases 124:S20–S25.

31. Oudghiri A, Karimi H, Chetioui F, Zakham F, Bourkadi JE, Elmessaoudi MD, Laglaoui A, Chaoui I, El Mzibri M. 2018. Molecular characterization of mutations associated with resistance to second-line tuberculosis drug among multidrug-resistant tuberculosis patients from high prevalence tuberculosis city in Morocco. BMC Infect Dis 18:98.

32. Momen G, Aainouss A, Lamaammal A, Chettioui F, Blaghen M, Messoudi M, Belghmi K, Mouslim J, El Mzibri M, El Messaoudi MD. 2021. Molecular characterization of mutations associated with resistance to second line drugs in Mycobacterium tuberculosis patients from Casablanca, Morocco. Rev Inst Med Trop Sao Paulo 63:e19.

33. Centner CM, Munir R, Tagliani E, Rieß F, Brown P, Hayes C, Dolby T, Zemanay W, Cirillo DM, David A. 2024. Reflex Xpert MTB/XDR testing of residual rifampicin-resistant specimens: a clinical laboratory-based diagnostic accuracy and feasibility study in South Africa, p. ofae437. *In* Open Forum Infectious Diseases. Oxford University Press US.

34. Pfurtscheller T, Tsutsunava A, Maghradze N, Gujabidze M, Bablishvili N, Yerlikaya S, Denkinger CM, Tukvadze N, Gupta-Wright A. 2025. Programmatic diagnostic accuracy and clinical utility of Xpert MTB/XDR in patients with rifampicin-resistant tuberculosis in Georgia, p. ofaf022. *In* Open Forum Infectious Diseases. Oxford University Press US.

35. Penn-Nicholson A, Gomathi SN, Ugarte-Gil C, Meaza A, Lavu E, Patel P, Choudhury B, Rodrigues C, Chadha S, Kazi M. 2021. A prospective multicentre diagnostic accuracy study for the Truenat tuberculosis assays. European Respiratory Journal 58:2100526.

36. Pillay S, Davies GR, Chaplin M, De Vos M, Schumacher SG, Warren R, Steingart KR, Theron G. 2021. Xpert MTB/XDR for detection of pulmonary tuberculosis and resistance to isoniazid, fluoroquinolones, ethionamide, and amikacin. Cochrane Database of Systematic Reviews 10.1002/14651858.CD014841.

37. Viney K, Linh NN, Gegia M, Zignol M, Glaziou P, Ismail N, Kasaeva T, Mirzayev F. 2021. New definitions of pre-extensively and extensively drug-resistant tuberculosis: update from the World Health Organization. European Respiratory Journal. European Respiratory Society.

38. Roelens M, Battista Migliori G, Rozanova L, Estill J, Campbell JR, Cegielski JP, Tiberi S, Palmero D, Fox GJ, Guglielmetti L. 2021. Evidence-based definition for extensively drug-resistant tuberculosis. Am J Respir Crit Care Med 204:713–722.

39. Dookie N, Rambaran S, Padayatchi N, Mahomed S, Naidoo K. 2018. Evolution of drug resistance in Mycobacterium tuberculosis: a review on the molecular determinants of resistance and implications for personalized care. Journal of Antimicrobial Chemotherapy 73:1138–1151.

40. Ennassiri W, Jaouhari S, Cherki W, Charof R, Filali-Maltouf A, Lahlou O. 2017. Extensively drug-resistant tuberculosis (XDR-TB) in Morocco. J Glob Antimicrob Resist 11:75–80.

41. Telenti A, Imboden P, Marchesi F, Matter L, Schopfer K, Bodmer T, Lowrie D, Colston MJ, Cole S. 1993. Detection of rifampicin-resistance mutations in Mycobacterium tuberculosis. The Lancet 341:647–651.

42. Miotto P, Tessema B, Tagliani E, Chindelevitch L, Starks AM, Emerson C, Hanna D, Kim PS, Liwski R, Zignol M. 2017. A standardised method for interpreting the association between mutations and phenotypic drug resistance in Mycobacterium tuberculosis. European Respiratory Journal 50:1701354.

43. Walker TM, Miotto P, Köser CU, Fowler PW, Knaggs J, Iqbal Z, Hunt M, Chindelevitch L, Farhat MR, Cirillo DM. 2022. The 2021 WHO catalogue of Mycobacterium tuberculosis complex mutations associated with drug resistance: a genotypic analysis. Lancet Microbe 3:e265–e273.

44. Van Deun A, Aung KJ, Hossain A, De Rijk P, Gumusboga M, Rigouts L, De Jong BC. 2015. Disputed rpoB mutations can frequently cause important rifampicin resistance among new tuberculosis patients.

45. Lin W-H, Lee W-T, Tsai H-Y, Jou R. 2021. Disputed rpoB mutations in Mycobacterium tuberculosis and tuberculosis treatment outcomes. Antimicrob Agents Chemother 65:10–1128.

46. Finci I, Albertini A, Merker M, Andres S, Bablishvili N, Barilar I, Cáceres T, Crudu V, Gotuzzo E, Hapeela N. 2022. Investigating resistance in clinical Mycobacterium tuberculosis complex isolates with genomic and phenotypic antimicrobial susceptibility testing: a multicentre observational study. Lancet Microbe 3:e672–e682.

47. Heyckendorf J, Andres S, Köser CU, Olaru ID, Schön T, Sturegård E, Beckert P, Schleusener V, Kohl TA, Hillemann D. 2018. What is resistance? Impact of phenotypic versus molecular drug resistance testing on therapy for multi-and extensively drug-resistant tuberculosis. Antimicrob Agents Chemother 62:10–1128.

48. Vilchèze C, Jacobs Jr WR. 2014. Resistance to isoniazid and ethionamide in Mycobacterium tuberculosis: genes, mutations, and causalities. Microbiol Spectr 2:10– 1128.

49. Seifert M, Catanzaro D, Catanzaro A, Rodwell TC. 2015. Genetic mutations associated with isoniazid resistance in Mycobacterium tuberculosis: a systematic review. PLoS One 10:e0119628.

50. Amri A, Benaissa E, Benlahlou Y, Bsaibis F, Maleb A, Chadli M, Elouenass M. 2025. Genotypic study of isolated resistance to isoniazid in the Mycobacterium tuberculosis complex in a Moroccan hospital. Access Microbiol 7:000928–v5.

51. Banerjee A, Dubnau E, Quemard A, Balasubramanian V, Um KS, Wilson T, Collins D, De Lisle G, Jacobs Jr WR. 1994. inhA, a gene encoding a target for isoniazid and ethionamide in Mycobacterium tuberculosis. Science (1979) 263:227–230.

52. DeBarber AE, Mdluli K, Bosman M, Bekker L-G, Barry 3rd CE. 2000. Ethionamide activation and sensitivity in multidrug-resistant Mycobacterium tuberculosis. Proceedings of the National Academy of Sciences 97:9677–9682.

53. Baulard AR, Betts JC, Engohang-Ndong J, Quan S, McAdam RA, Brennan PJ, Locht C, Besra GS. 2000. Activation of the pro-drug ethionamide is regulated in mycobacteria. Journal of Biological Chemistry 275:28326–28331.

54. Morlock GP, Metchock B, Sikes D, Crawford JT, Cooksey RC. 2003. ethA, inhA, and katG loci of ethionamide-resistant clinical Mycobacterium tuberculosis isolates. Antimicrob Agents Chemother 47:3799–3805.

55. Malik S, Willby M, Sikes D, Tsodikov O V, Posey JE. 2012. New insights into fluoroquinolone resistance in Mycobacterium tuberculosis: functional genetic analysis of gyrA and gyrB mutations. PLoS One 7:e39754.

56. Avalos E, Catanzaro D, Catanzaro A, Ganiats T, Brodine S, Alcaraz J, Rodwell T. 2015. Frequency and geographic distribution of gyrA and gyrB mutations associated with fluoroquinolone resistance in clinical Mycobacterium tuberculosis isolates: a systematic review. PLoS One 10:e0120470.

57. Maruri F, Sterling TR, Kaiga AW, Blackman A, van der Heijden YF, Mayer C, Cambau E, Aubry A. 2012. A systematic review of gyrase mutations associated with fluoroquinolone-resistant Mycobacterium tuberculosis and a proposed gyrase numbering system. Journal of Antimicrobial Chemotherapy 67:819–831.

58. Farhat MR, Sultana R, Iartchouk O, Bozeman S, Galagan J, Sisk P, Stolte C, Nebenzahl-Guimaraes H, Jacobson K, Sloutsky A. 2016. Genetic determinants of drug resistance in Mycobacterium tuberculosis and their diagnostic value. Am J Respir Crit Care Med 194:621–630.

59. Chen X, Li R, Ge S, Li Y, Cai C, Weng T, Zhang Y, Jiang J, Feng Z, Chen Y. 2023. Rapid detection of extensive drug resistance by Xpert MTB/XDR optimizes therapeutic decision-making in rifampin-resistant tuberculosis patients. J Clin Microbiol 61:e01832–22.

60. Georghiou SB, Penn-Nicholson A, de Vos M, Mace A, Syrmis MW, Jacob K, Mape A, Parmar H, Cao Y, Coulter C. 2021. Analytical performance of the Xpert MTB/XDR® assay for tuberculosis and expanded resistance detection. Diagn Microbiol Infect Dis 101:115397.

61. Engelthaler DM, Streicher EM, Kelley EJ, Allender CJ, Wiggins K, Jimenez D, Lemmer D, Vittinghoff E, Theron G, Sirgel FA. 2019. Minority Mycobacterium tuberculosis genotypic populations as an indicator of subsequent phenotypic resistance. Am J Respir Cell Mol Biol 61:789–791.

62. Tolani MP, D’souza DTB, Mistry NF. 2012. Drug resistance mutations and heteroresistance detected using the GenoType MTBDR plus assay and their implication for treatment outcomes in patients from Mumbai, India. BMC Infect Dis 12:9.

63. Cuella-Martin I, Hakizayezu F, Ahmed A, Runyambo D, Niyompano H, Keysers J, De Rijk WB, Mulders W, Mitchell EMH, Decroo T. 2025. Paucibacillary tuberculosis drives the low positive predictive value of Xpert MTB/RIF Ultra for rifampicin resistance detection in low-prevalence settings. Clinical Infectious Diseases 81:372– 378.

64. Hossain A, Asmar A El, Almheiri A, Almheiri H, Alameddine M. 2026. Global and regional trends in multidrug-or rifampicin-resistant tuberculosis, 2000–2024 with forecasts to 2030: a systematic review and meta-analysis. BMC Infect Dis.

65. McQuaid CF, Horton KC, Dean AS, Knight GM, White RG. 2020. The risk of multidrug-or rifampicin-resistance in males versus females with tuberculosis. European Respiratory Journal 56:2000626.

66. Rahman SMM, Ruhee NN, Haider A, Hasan MJ, Nasrin R, Adel ASS, Uddin MKM, Ahmed S, Maug AKJ, Banu S. 2025. Xpert MTB/XDR Assay for Detection of Resistance to Isoniazid, Fluoroquinolone, Aminoglycoside, and Ethionamide Among Patients with Pulmonary Tuberculosis in Bangladesh. Infect Dis Ther 14:803–818.

67. Omar SV, Louw G, Ismail F, Liu X, Ngcamu D, Gwala T, van der Meulen M, Joseph L. 2024. Performance evaluation of the Xpert MTB/XDR test for the detection of drug resistance to Mycobacterium tuberculosis among people diagnosed with tuberculosis in South Africa. J Clin Microbiol 62:e00229–24.

68. Keller S, Naidoo K, Zekarias M, Israel-Isah S, Shaka M, Gule G, Naidoo A, Bathnna M, Dlamini-Miti JN, Yae K. 2025. Feasibility and acceptability of GeneXpert MTB/XDR implementation among healthcare workers in three low-middle income African countries. PLoS One 20:e0326342.

69. Saukkonen JJ, Duarte R, Munsiff SS, Winston CA, Mammen MJ, Abubakar I, Acuña-Villaorduña C, Barry PM, Bastos ML, Carr W. 2025. Updates on the treatment of drug-susceptible and drug-resistant tuberculosis: an official ATS/CDC/ERS/IDSA clinical practice guideline. Am J Respir Crit Care Med 211:15–33.

70. Schwab TC, Perrig L, Göller PC, De la Hoz FFG, Lahousse AP, Minder B, Günther G, Efthimiou O, Omar SV, Egger M. 2024. Targeted next-generation sequencing to diagnose drug-resistant tuberculosis: a systematic review and meta-analysis. Lancet Infect Dis 24:1162–1176.

71. Chaoui I, Oudghiri A, Chetioui F, Laglaoui A, El Messaoudi MD. 2018. GENOTYPING OF MULTIDRUG AND PREEXTENSIVELY DRUG-RESISTANT MYCOBACTERIUM TUBERCULOSIS ISOLATES FROM A HIGH INCIDENCE TB AREA IN MOROCCO. Russian Journal of Infection and Immunity 8:565.

72. Oudghiri A, Momen G, Aainouss A, Laglaoui A, El Messaoudi MD, El Mzibri M, Chaoui I. 2021. Genotypic diversity of multi-and pre-extremely drug-resistant Mycobacterium tuberculosis isolates from Morocco. PLoS One 16:e0253826.

73. Casali N, Nikolayevskyy V, Balabanova Y, Harris SR, Ignatyeva O, Kontsevaya I, Corander J, Bryant J, Parkhill J, Nejentsev S. 2014. Evolution and transmission of drug-resistant tuberculosis in a Russian population. Nat Genet 46:279–286.

74. Wollenberg KR, Desjardins CA, Zalutskaya A, Slodovnikova V, Oler AJ, Quiñones M, Abeel T, Chapman SB, Tartakovsky M, Gabrielian A. 2017. Whole-genome sequencing of Mycobacterium tuberculosis provides insight into the evolution and genetic composition of drug-resistant tuberculosis in Belarus. J Clin Microbiol 55:457–469.

75. Xie YL, Chakravorty S, Armstrong DT, Hall SL, Via LE, Song T, Yuan X, Mo X, Zhu H, Xu P. 2017. Evaluation of a rapid molecular drug-susceptibility test for tuberculosis. New England Journal of Medicine 377:1043–1054.

