## Supplemental File 2 for "Routine Xpert MTB/XDR for Tuberculosis Drug-Resistance Surveillance in Northern Morocco: A Four-Year Programmatic Analysis"

**Supplementary Table S2. Complete melting-temperature analysis of wild-type and mutation-associated signals across Xpert MTB/XDR and MTB/RIF *Ultra* targets|** Melting temperatures (Tm) are presented as median (interquartile range [IQR]) for wild-type (WT) and mutation-associated signals across resistance-associated targets from Xpert MTB/XDR and linked Xpert MTB/RIF Ultra results. The analysis includes the INH-associated targets ***inhA, katG* and *ahpC*,** the RIF-associated ***rpoB1, rpoB2, rpoB3* and *rpoB4-A*** targets, and the FQ-associated ***gyrA1, gyrA2* and *gyrA3*** targets. Differences represent the mutation-associated median Tm minus the WT median Tm.

| **Target** | **WT n** | **WT median (IQR), °C** | **Mutation n** | **Mutation median (IQR), °C** | **Difference, °C** | **Holm-adjusted P** |
| --- | --- | --- | --- | --- | --- | --- |
| ***inhA*** | 173 | 76.2 (76.1–76.3) | 26 | 70.8 (70.7–70.9) | −5.40 | <0.00001 |
| ***katG*** | 150 | 73.7 (73.6–73.7) | 38 | 68.2 (68.1–68.6) | −5.50 | <0.00001 |
| ***ahpC*** | 166 | 68.6 (68.5–68.7) | 14 | 66.0 (65.9–66.2) | −2.60 | <0.00001 |
| ***rpoB1*** | 169 | 69.3 (69.2–69.4) | 15 | 65.5 (64.0–65.7) | −3.80 | <0.00001 |
| ***rpoB2*** | 168 | 73.1 (73.0–73.2) | 24 | 69.5 (69.1–70.2) | −3.60 | <0.00001 |
| ***rpoB3*** | 144 | 75.7 (75.5–75.8) | 46 | 73.0 (72.1–73.3) | −2.70 | <0.00001 |
| ***rpoB4-A*** | 114 | 67.0 (66.9–67.1) | 43 | 73.5 (73.3–73.7) | +6.50 | <0.00001 |
| ***gyrA1*** | 174 | 76.3 (76.2–76.4) | 18 | 72.0 (71.9–72.5) | −4.25 | <0.00001 |
| ***gyrA2*** | 174 | 70.2 (70.1–70.3) | 13 | 75.6 (75.4–75.7) | +5.40 | <0.00001 |
| ***gyrA3*** | 175 | 71.0 (71.0–71.1) | 17 | 76.2 (69.7–76.3) | +5.20 | 0.00426 |

Analysis was restricted to MTB-detected testing episodes with numeric Tm measurements. One numeric mutation-associated Tm value per target per testing episode was retained across Xpert MTB/XDR and MTB/RIF*Ultra* targets. Comparisons between WT and mutation-associated Tm distributions were performed using two-sided Mann–Whitney U tests, with P values adjusted for multiple comparisons using the Holm method across the 10 analyzable target-level comparisons. *fabG1*, *gyrB2* and *eis* had no numeric mutation-associated Tm observations and were therefore not included in inferential analysis; *rrs* had only one mutation-associated observation and was excluded from inferential comparison. Tm patterns represent assay-level molecular signals and should not be interpreted as identification of specific nucleotide substitutions.
