## Supplemental File 3 for "Routine Xpert MTB/XDR for Tuberculosis Drug-Resistance Surveillance in Northern Morocco: A Four-Year Programmatic Analysis"

**Supplementary table S3. Melt-peak height characteristics of wild-type and mutation-associated signals across resistance-associated targets |** Peak heights are presented as median (interquartile range [IQR]) for wild-type (WT) and mutation-associated signals. The analysis was restricted to MTB-detected testing episodes with numeric peak-height measurements. WT peak heights were derived from observations without a numeric mutation-associated signal for the corresponding target, whereas all available numeric mutation-associated peak heights were retained. Differences between WT and mutation-associated peak-height distributions were assessed using two-sided Mann–Whitney U tests. P values were adjusted for multiple comparisons using the Holm method across the 10 analyzable target-level comparisons.

| **Drug class** | **Target** | **WT n** | **WT peak height, median (IQR)** | **Mutation-associated (n)** | **Mutation-associated peak height, median (IQR)** | **Holm-adjusted *P*** |
| --- | --- | --- | --- | --- | --- | --- |
| INH | *inhA* | 173 | 233.5 (217.1–248.4) | 26 | 233.7 (207.6–256.5) | 0.623 |
| INH | *katG* | 150 | 80.4 (71.8–87.5) | 38 | **121.0 (112.2–135.6)** | **<0.0001** |
| INH | *ahpC* | 166 | 37.8 (33.3–42.5) | 14 | **45.0 (39.2–52.0)** | **0.048** |
| FQ | *gyrA1* | 174 | 44.2 (39.0–49.7) | 18 | **63.8 (41.9–76.8)** | **0.037** |
| FQ | *gyrA2* | 174 | 64.2 (53.8–73.2) | 13 | **138.4 (118.1–163.4)** | **<0.0001** |
| FQ | *gyrA3* | 175 | 72.3 (62.4–79.0) | 17 | 64.0 (56.7–75.6) | 0.346 |
| Injectable | *rrs* | 195 | 165.5 (149.4–178.8) | 1 | 30.7 (30.7–30.7) | — |
| RIF | *rpoB1* | 170 | 58.5 (53.4–68.1) | 15 | **38.5 (33.5–59.2)** | **0.006** |
| RIF | *rpoB2* | 168 | 72.7 (65.7–88.2) | 24 | 39.1 (20.2–86.4) | 0.346 |
| RIF | *rpoB3* | 144 | 73.0 (65.2–85.0) | 46 | **37.5 (29.4–44.6)** | **<0.0001** |
| RIF | *rpoB4* | 114 | 54.0 (48.6–59.5) | 43 | **110.6 (110.2–117.1)** | **<0.0001** |

* The rrs mutation-associated group contained only one observation (n=1) and is therefore descriptive only; the reported P value should not be interpreted as an inferential comparison.
† fabG1, gyrB2, and eis had no numeric mutation-associated peak-height observations and were therefore excluded from inferential comparisons. Peak-height differences describe **assay-level signal characteristics** and should not be interpreted as evidence of specific nucleotide substitutions. **Abbreviations:** MTB, Mycobacterium tuberculosis; Xpert MTB/XDR, Xpert MTB/XDR assay; WT, wild type; IQR, interquartile range; INH, isoniazid; FQ, fluoroquinolones; RIF, rifampicin.
