## Supplemental File 4 for "Routine Xpert MTB/XDR for Tuberculosis Drug-Resistance Surveillance in Northern Morocco: A Four-Year Programmatic Analysis"

**Supplementary Table S4. Associations between demographic characteristics and molecular resistance profiles|** Associations between demographic characteristics and selected molecular resistance outcomes were assessed among episodes with interpretable results for the corresponding outcome. For sex and province, values in the predictor column are presented as outcome-positive episodes/total episodes within each category. For age, values represent the median age among outcome-positive versus outcome-negative episodes. Odds ratios (ORs) compare males with females. Province comparisons represent global tests across all provincial categories; no single OR is applicable. P values are unadjusted, and FDR-adjusted P values were calculated using the Benjamini–Hochberg procedure across the 18 prespecified demographic comparisons.

| **Outcome** | **Predictor** | **N** | **Effect / comparison** | **95% CI** | **P** | **FDR-adjusted P** |
| --- | --- | --- | --- | --- | --- | --- |
| RIF resistance | Sex: male 67/148  female 8/40 | 188 | OR 3.31 | 1.43–7.66 | 0.0036 | 0.0328 |
| RIF resistance | Age (continuous): resistant median 39.0 vs susceptible 41.5 years | 185 | Median 39.0 vs 41.5 | — | 0.2108 | 0.3163 |
| RIF resistance | Province: Tétouan 17/57; Tangier 27/53;  Al Hoceima 11/27; Ouazzane 7/18;  Fnideq 3/15;  Chefchaouen 7/11;  Larache 3/7. | 188 | χ²=10.21 | — | 0.1143 | 0.2802 |
| INH resistance | Sex: male 60/153  female 11/39 | 192 | OR 1.64 | 0.76–3.54 | 0.2652 | 0.3672 |
| INH resistance | Age (continuous): resistant median 38.0 vs susceptible 42.0 years | 190 | Median 38.0 vs 42.0 | — | 0.1558 | 0.2802 |
| INH resistance | Province: Tétouan 24/61; Tangier 25/55;  Al Hoceima 5/27;  Ouazzane 6/17;  Fnideq 1/14;  Chefchaouen 7/11;  Larache 3/7 | 192 | χ²=14.62 | — | 0.0207 | 0.1244 |
| FQ resistance | Sex: male 16/155  female 4/38 | 193 | OR 0.98 | 0.31–3.12 | 1.0000 | 1.0000 |
| FQ resistance | Age (continuous): resistant median 38.5 vs susceptible 41.0 years | 191 | Median 38.5 vs 41.0 | — | 0.3411 | 0.4093 |
| FQ resistance | Province: Tétouan 12/65; Tangier 2/52;  Al Hoceima 3/27;  Ouazzane 1/17;  Fnideq 1/14;  Chefchaouen 0/11;  Larache 1/7 | 193 | χ²=8.89 | — | 0.1712 | 0.2802 |
| **Outcome** | **Predictor** | **N** | **Effect / comparison** | **95% CI** | **P** | **FDR-adjusted P** |
| ETH resistance | Sex: male 20/160  female 6/40 | 200 | OR 0.81 | 0.30–2.17 | 0.6104 | 0.6463 |
| ETH resistance | Age (continuous): resistant median 30.5 vs susceptible 39.0 years | 197 | Median 30.5 vs 39.0 | — | 0.1553 | 0.2802 |
| ETH resistance | Province: Tétouan 3/67; Tangier 13/55;  Al Hoceima 0/27;  Ouazzane 4/18;  Fnideq 0/15;  Chefchaouen 5/11;  Larache 1/7 | 200 | χ²=27.69 | — | 0.0004 | 0.0072 |
| MDR profile | Sex: male 43/145  female 8/39 | 184 | OR 1.63 | 0.69–3.84 | 0.3163 | 0.4066 |
| MDR profile | Age (continuous): MDR median 38.0 vs non-MDR 41.0 years | 182 | Median 38.0 vs 41.0 | — | 0.0959 | 0.2802 |
| MDR profile | Province: Tétouan 14/57; Tangier 17/51;  Al Hoceima 5/27;  Ouazzane 5/17;  Fnideq 1/14;  Chefchaouen 7/11;  Larache 2/7 | 184 | χ²=12.30 | — | 0.0524 | 0.2360 |
| ≥1 additional resistance beyond RIF | Sex: male 61/146 vs female 14/40 | 186 | OR 1.33 | 0.64–2.76 | 0.4722 | 0.5312 |
| ≥1 additional resistance beyond RIF | Age (continuous): additional resistance median 38.0 vs none 43.0 years | 183 | Median 38.0 vs 43.0 | — | 0.1321 | 0.2802 |
| ≥1 additional resistance beyond RIF | Province: Tétouan 25/57; Tangier 23/51;  Al Hoceima 8/27;  Ouazzane 7/18;  Fnideq 2/15;  Chefchaouen 7/11;  Larache 3/7 | 186 | χ²=9.12 | — | 0.1676 | 0.2802 |

For age, the effect column gives the median age in resistant versus non-resistant episodes. For province, the effect column gives the global Pearson χ² statistic and no single odds ratio is reported because province has multiple categories.

Abbreviations: AMK, amikacin; CAP, capreomycin; ETH, ethionamide; FQ, fluoroquinolones; INH, isoniazid; KAN, kanamycin; MDR, multidrug-resistant; MTB, Mycobacterium tuberculosis complex; RIF, rifampicin; Tm, melting temperature; WT, wild type; FDR, false discovery rate.
